# Reduced antibody activity against SARS-CoV-2 B.1.617.2 Delta virus in serum of mRNA-vaccinated patients receiving TNF-α inhibitors

**DOI:** 10.1101/2021.09.28.21264250

**Authors:** Rita E. Chen, Matthew J. Gorman, Daniel Y. Zhu, Juan Manuel Carreño, Dansu Yuan, Laura A. VanBlargan, Samantha Burdess, Douglas A. Lauffenburger, Wooseob Kim, Jackson S. Turner, Lindsay Droit, Scott A. Handley, Salim Chahin, Parakkal Deepak, Jane A. O’Halloran, Michael Paley, Rachel M. Presti, Gregory F. Wu, Florian Krammer, Galit Alter, Ali H. Ellebedy, Alfred H. J. Kim, Michael S. Diamond

## Abstract

Although vaccines effectively prevent COVID-19 in healthy individuals, they appear less immunogenic in individuals with chronic inflammatory diseases (CID) and/or under chronic immunosuppression, and there is uncertainty of their activity against emerging variants of concern in this population. Here, we assessed a cohort of 74 CID patients treated as monotherapy with chronic immunosuppressive drugs for functional antibody responses in serum against historical and variant SARS-CoV-2 viruses after immunization with Pfizer mRNA BNT162b2 vaccine. Longitudinal analysis showed the greatest reductions in neutralizing antibodies and Fc effector function capacity in individuals treated with TNF-α inhibitors, and this pattern appeared worse against the B.1.617.2 Delta virus. Within five months of vaccination, serum neutralizing titers of the majority of CID patients fell below the presumed threshold correlate for antibody-mediated protection. Thus, further vaccine boosting or administration of long-acting prophylaxis (*e*.*g*., monoclonal antibodies) likely will be required to prevent SARS-CoV-2 infection in this susceptible population.

## INTRODUCTION

In December 2019, severe acute respiratory syndrome coronavirus 2 (SARS-CoV-2) emerged and the global COVID-19 pandemic began. Since then, many antibody-based therapeutics and vaccines have been developed (Case et al., 2021; Sempowski et al., 2020) with some given Emergency Use Authorization (EUA) or Food and Drug Administration approval (*e*.*g*., BNT162b2 mRNA vaccine) in hopes of preventing infection and severe disease. While several of these countermeasures show efficacy against historical (2019-early 2020) SARS-CoV-2 strains, the emergence of variants of concern (VOC) has prompted questions as to whether they will retain efficacy.

SARS-CoV-2 spike protein engages cell-surface receptor angiotensin-converting enzyme 2 (ACE2) for attachment and entry into human cells (Letko et al., 2020). The S1 component of the spike protein contains the N-terminal (NTD) and receptor binding (RBD) domains, the latter being the primary target of neutralizing antibodies (Cao et al., 2020; Pinto et al., 2020; Shi et al., 2020; Zost et al., 2020). However, recent studies have shown that therapeutic monoclonal and vaccine-elicited polyclonal antibodies have reduced neutralizing activity against VOC, likely because these strains contain mutations within the RBD and the receptor binding motif (RBM) (Chen et al., 2021a; Chen et al., 2021b; Liu et al., 2021b; Liu et al., 2021c; Wang et al., 2021b). This observation is concerning since serum neutralizing antibody titers are believed to be an *in vitro* correlate of *in vivo* protection (Corbett et al., 2021a; Corbett et al., 2021b; Francica et al., 2021).

Most studies on the immunogenicity and efficacy of SARS-CoV-2 vaccines have focused on immunocompetent animals and humans. As SARS-CoV-2 has been documented to mutate and evolve in immunocompromised hosts (Choi et al., 2020; Clark et al., 2021), it is important to understand whether vaccine-elicited responses are protective and durable in this population. Just recently, the Centers for Disease Control and Prevention recommended an additional BNT162b2 mRNA dose for the immunocompromised notwithstanding the relatively limited study of effects of immunosuppression on the immunogenicity and protection following SARS-CoV-2 vaccination. Here, we examine a cohort of chronic inflammatory disease (CID) patients from the COVID-19 Vaccine Responses in Patients with Autoimmune Disease (COVaRiPAD) study (Deepak et al., 2021) receiving different treatment regimens for functional antibody responses against SARS-CoV-2 at three- and five-months after completion of a two-dose BNT162b2 mRNA vaccination regimen.

## RESULTS

Although the potency of neutralization by monoclonal antibodies (mAbs) can be affected greatly by small numbers of mutations in emerging SARS-CoV-2 strains (Chen et al., 2021a; Chen et al., 2021b; Planas et al., 2021; Wang et al., 2021a; Wang et al., 2021b), it remains less clear how polyclonal antibodies derived after vaccination that target multiple epitopes perform. Whereas the efficacy of mRNA vaccine protection in immunocompetent volunteers has been high (>90%) against historical and some emerging SARS-CoV-2 strains (*e*.*g*., B.1.1.7 [Alpha]) (Abu-Raddad et al., 2021; Chemaitelly et al., 2021), there are concerns for breakthrough infections in immunosuppressed individuals or those receiving immunomodulatory drugs because of blunted immune responses, waning immunity, or evasion by emerging variants including B.1.351 (Beta) and B.1.617.2 (Delta). We assessed the neutralizing activity of serum from individuals immunized with the BNT162b2 mRNA vaccine against fully infectious SARS-CoV-2 strains including recombinant WA1/2020 viruses encoding D614G (WA1/2020 D614G) or the B.1.351 (Beta) spike (Wash-B.1.351) and a clinical isolate of B.1.617.2 (Delta). All viruses were passaged in Vero-TMPRSS2 cells to minimize adventitious generation of furin cleavage site mutations (Klimstra et al., 2020) and confirmed by deep sequencing (**Table S1**).

We obtained sera from a cohort of CID patients 3 months after BNT162b2 mRNA vaccination. This group included patients with Crohn’s disease, ulcerative colitis, asthma, multiple sclerosis, ankylosing spondylitis, systemic lupus erythematosus, Sjögren’s syndrome, Hashimoto’s disease, psoriasis, rheumatoid arthritis and undifferentiated inflammatory arthritis, type 1 diabetes, combined variable immune deficiency, alopecia areata, uveitis, vasculitis, scleroderma, psoriatic arthritis, anti-neutrophil cytoplasmic antibody (ANCA)-associated vasculitis, and microscopic colitis (**Table 1**). Because of their disease severity, some patients received multiple drug interventions. For our analysis, we focused on subjects (n = 74) treated with a single immunomodulatory agent so we could correlate treatment interventions with immunological outcomes. This included patients receiving chronic treatment with antimetabolites (n = 12), TNF-α inhibitors (TNFi) (n = 11), antimalarial agents (n = 10), anti-integrin inhibitors (n = 10), non-steroidal anti-inflammatory drugs (NSAIDs) (n = 9), anti-IL-23 inhibitor (n = 9), B cell depletion therapy (BCDT) (n = 5), Bruton’s tyrosine kinase inhibitor (BTKi) (n = 1), nuclear factor-erythroid factor 2-related factor 2 (Nrf2) activator (n = 1), sulfasalazine (SSZ) (n = 1), systemic steroids (n = 2), anti-B lymphocyte stimulator (anti-BLyS) (n=1), or sphingosine 1-phosphate receptor modulator (S1PR mod) (n = 1).

**Table 1.** Patient characteristics.

|  |  | **Total N=74** |
| --- | --- | --- |
|  |  | **N (%)** |
| **Age (median [range])** |  | 48 (22-82) |
| **Sex** |  |  |
|  | Female | 52 (70) |
|  | Male | 21 (28) |
|  | Other | 1 (1) |
| **Race** |  |  |
|  | White | 66 (89) |
|  | Black | 5 (7) |
|  | Multiple Race | 2 (3) |
|  | Asian | 1 (1) |
| **Chronic inflammatory disease<sup>a</sup>** |  |  |
|  | Alopecia Areata | 1 (1) |
|  | ANCA Associated Vasculitis | 1 (1) |
|  | Ankylosing Spondylitis | 2 (3) |
|  | Asthma | 6 (8) |
|  | Combined Variable Immune Deficiency | 2 (3) |
|  | Crohn's Disease | 31 (42) |
|  | Hashimoto's Disease | 5 (7) |
|  | IBD Associated Arthritis | 1 (1) |
|  | Inflammatory Arthritis | 1 (1) |
|  | Multiple Sclerosis | 10 (14) |
|  | Psoriasis | 4 (5) |
|  | Psoriatic Arthritis | 1 (1) |
|  | Rheumatoid Arthritis | 8 (11) |
|  | Scleroderma | 1 (1) |
|  | Sjogren's Syndrome | 5 (7) |
|  | Systemic Lupus Erythematosus | 5 (7) |
|  | Type 1 Diabetes | 4 (5) |
|  | Ulcerative Colitis | 5 (7) |
|  | Uveitis | 1 (1) |
|  | Other | 2 (3) |

<sup>a</sup> Patients may have more than one disease diagnosis.

We first used an ELISA to assess serum IgG binding to spike proteins of Wuhan-1 (historical) or three dominant circulating variants: B.1.1.7, B.1.351, and B.1.617.2. When analyzed for relative binding to the different spike proteins, sera from individual immunocompetent, healthy volunteers (n = 25) (Turner et al., 2021), CID patients (n = 45), or CID patients treated with antimalarial monotherapy showed slightly reduced (1.5-fold) binding to B.1.1.7, B.1.351, and B.1.617.2 than to Wuhan-1 spike protein (**Fig 1A-B and E**). Serum IgG titers from CID patients as a group trended lower against the historical and variant spike proteins than immunocompetent volunteers, although the differences did not attain statistical significance (WT: *p* = 0.6; B.1.1.7: *p* = 0.29; B.1.351: *p* = 0.14; B.1.617.2: *p* = 0.23) (**Fig 1A-B**). Anti-spike IgG binding titers of subjects receiving TNFi trended lower (2 to 3-fold) than immunocompetent controls (WT: *p* = 0.6; B.1.1.7: *p* = 0.48; B.1.351: *p* = 0.3; B.1.617.2: *p* = 0.4) (**Fig 1A and C**). Most other drug treatment groups with enough patients enrolled (n ≥ 3) did not show differences in anti-spike IgG binding titers compared to the controls (**Fig 1D-G**). Other treatment groups had too few patients enrolled to interpret trends, but as expected, patients receiving BCDT had low levels of IgG against the spike proteins (**Fig S1**).

**Figure 1.**
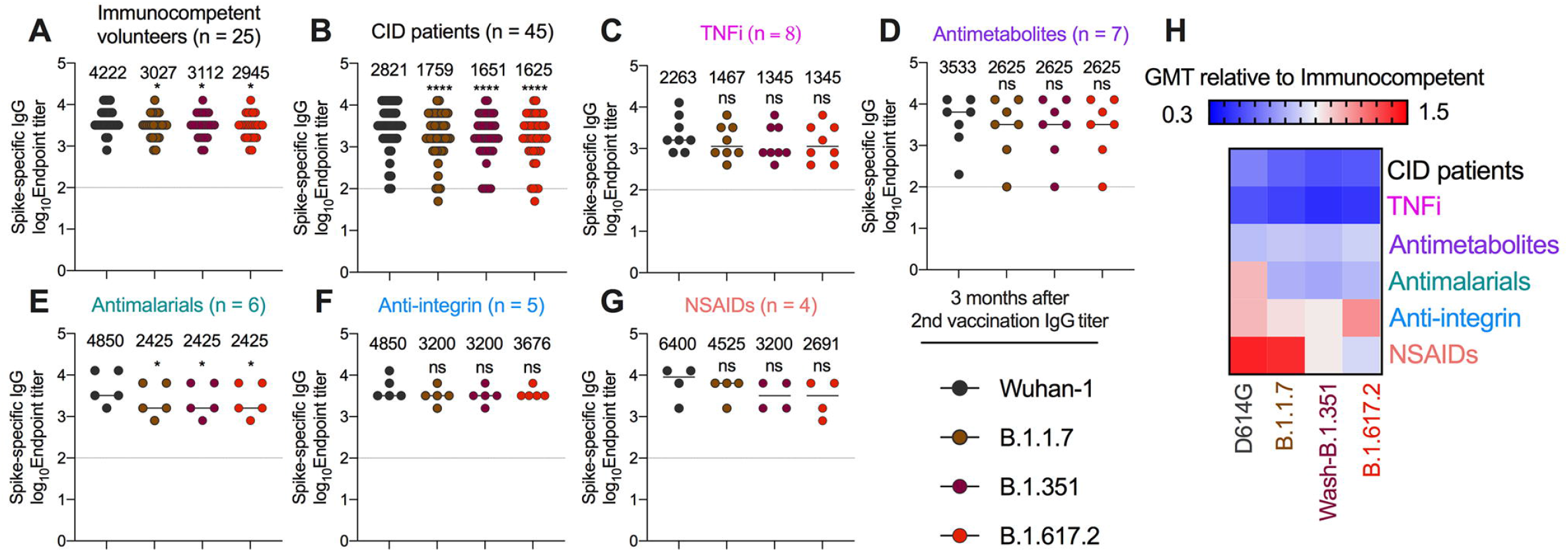
Serum IgG titers against SARS-CoV-2 variant spike proteins at three months after second vaccination. (**A-G**) Paired analyses of spike-specific endpoint IgG serum titers measured by ELISA from humans at three months after second vaccination with BNT162b2 mRNA vaccine. Individuals were grouped as (**A**) immunocompetent volunteers (n = 25) and (**B**) CID patients (n = 45) or subdivided by immunosuppressive drug class (**C**) TNFi (n = 8), (**D**) antimetabolites (n = 7), (**E**) antimalarials (n = 6), (**F**) anti-integrin inhibitors (n = 5), or (**G**) non-steroidal anti-inflammatory drugs (NSAIDs) (n = 4). Geometric mean titer (GMT) values are shown on graph. Dotted line represents limit of detection of the assay. One-way ANOVA with Dunnett’s post-test; * *p* < 0.05; **** *p* < 0.0001. (**H**) Heat map of GMT values relative to healthy volunteer GMT values for each SARS-CoV-2 spike protein. Blue, reduction; red, increase.

To begin to determine the functional capacity of the antibody response, we interrogated sera obtained from immunocompetent volunteers (n = 25) and CID patients (n = 74) three months after their second dose of BNT162b2 mRNA vaccine. As a group, sera from CID patients trended towards lower neutralizing activity, although comparisons did not attain statistical significance (WA1/2020 D614G (Geometric mean titer [GMT]: 239 [immunocompetent] versus 177 [CID patients], *p* = 0.98); Wash-B.1.351 (GMT: 117 [immunocompetent] versus 80 [CID patients], *p* = 0 .67); and B.1.617.2 (GMT: 109 [immunocompetent] versus 84 [CID patients], *p* = 0.98) (**Fig 2A-B**). In both control and CID patient groups, the B.1.351 and B.1.617.2 variants were neutralized less efficiently than the historical WA1/2020 D614G strain, as reported previously (Chen et al., 2021b; Liu et al., 2021a; Liu et al., 2021b; Planas et al., 2021; Wang et al., 2021a). Among the treatment subgroups, patients receiving TNFi had lower neutralizing titers (GMT: 239 (immunocompetent) versus 104 (TNFi) [WA1/2020 D614G], *p* = 0.26; 117 versus 46 [Wash-B.1.351] *p* = 0.04; 109 versus 50 [B.1.617.2], *p* = 0.15) (**Fig 2A and D**). Other treatment groups (n > 5) including antimalarial, anti-integrin, NSAIDs, and anti-IL-23 inhibitors had similar neutralization titers to immunocompetent volunteers (**Fig 2A and E-H**). In addition, all subgroups with n > 5 neutralized Wash-B.1.351 and B.1.617.2 less efficiently than WA1/2020 (**Fig 2A-H**). These trends in neutralization titers also were seen in groups receiving other immunosuppressive agents with smaller number of individuals (**Fig S2**).

**Figure 2.**
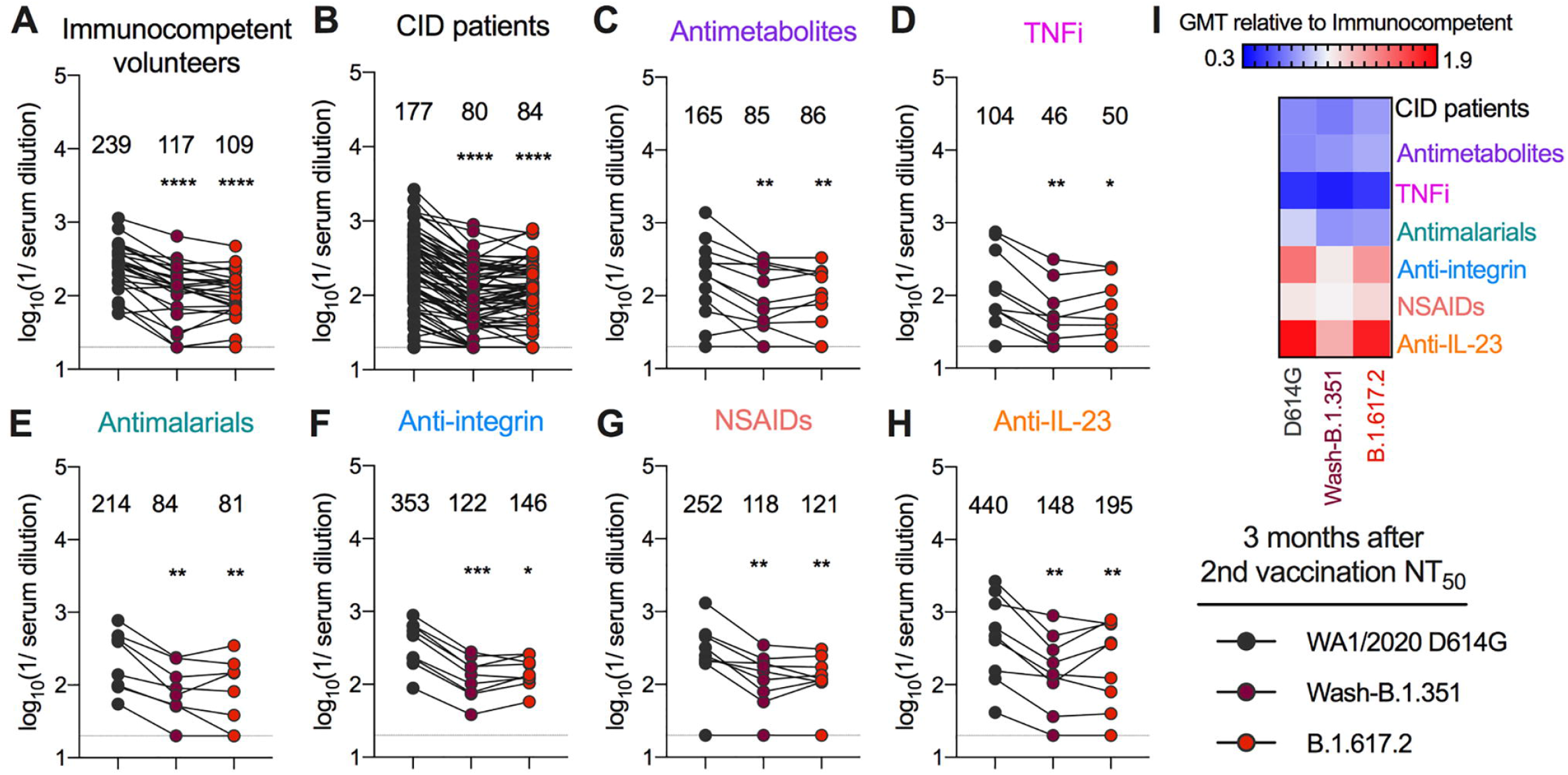
Serum neutralization titers against SARS-CoV-2 variant viruses at three months after second vaccination. (**A-H**) Paired analyses of neutralization (NT_50_) titers in serum measured by focus reduction neutralization test (FRNT) against fully infectious SARS-CoV-2 strains at three months after second vaccination with BNT162b2 mRNA vaccine. Individuals were grouped as (**A**) immunocompetent volunteers (n = 25) and (**B**) CID patients (n = 74) or subdivided by immunosuppressive drug class (**C**) antimetabolites (n = 12), (**D**) TNFi (n = 11), **(E)** antimalarials (n = 8), (**F**) anti-integrin inhibitors (n = 9), (**G**) NSAIDs (n = 9), or (**H**) anti-IL-23 inhibitors (n = 9). GMT values are shown on graph. Dotted line represents limit of detection of the assay. One way ANOVA with Dunn’s post-test; * *p* < 0.05; ** *p* < 0.01; *** *p* < 0.001; **** *p* < 0.0001. (**I**) Heat map of GMT values relative to healthy volunteer GMT values for each SARS-CoV-2 spike protein. Blue, reduction; red, increase.

A recent study analyzing the relationship between *in vitro* neutralization titers and protection against SARS-CoV-2 infection by vaccines estimated a protective serum titer of 1/54 (range 1/10 to 1/200 depending on the assay used) (Khoury et al., 2021). Given this result, we determined the fraction of individuals in the different groups of our cohort that fell below this cut-off. Three months after the completion of the initial series of BNT162b2 mRNA vaccine, only 2 of 25 (8%) immunocompetent volunteers had neutralizing titers (NT_50_) below 1/50 against B.1.617.2, whereas 30 of 74 (35%) of CID patients had titers below this level (**Table 2**). Many of the drug treatment groups also had a higher percentage of individuals below the 1/50 cut-off than immunocompetent volunteers including antimetabolites (33%), TNFi (64%), antimalarial agents (30%), NSAIDs (11%), and anti-IL-23 inhibitor (22%) (**Table 2**).

**Table 2.**
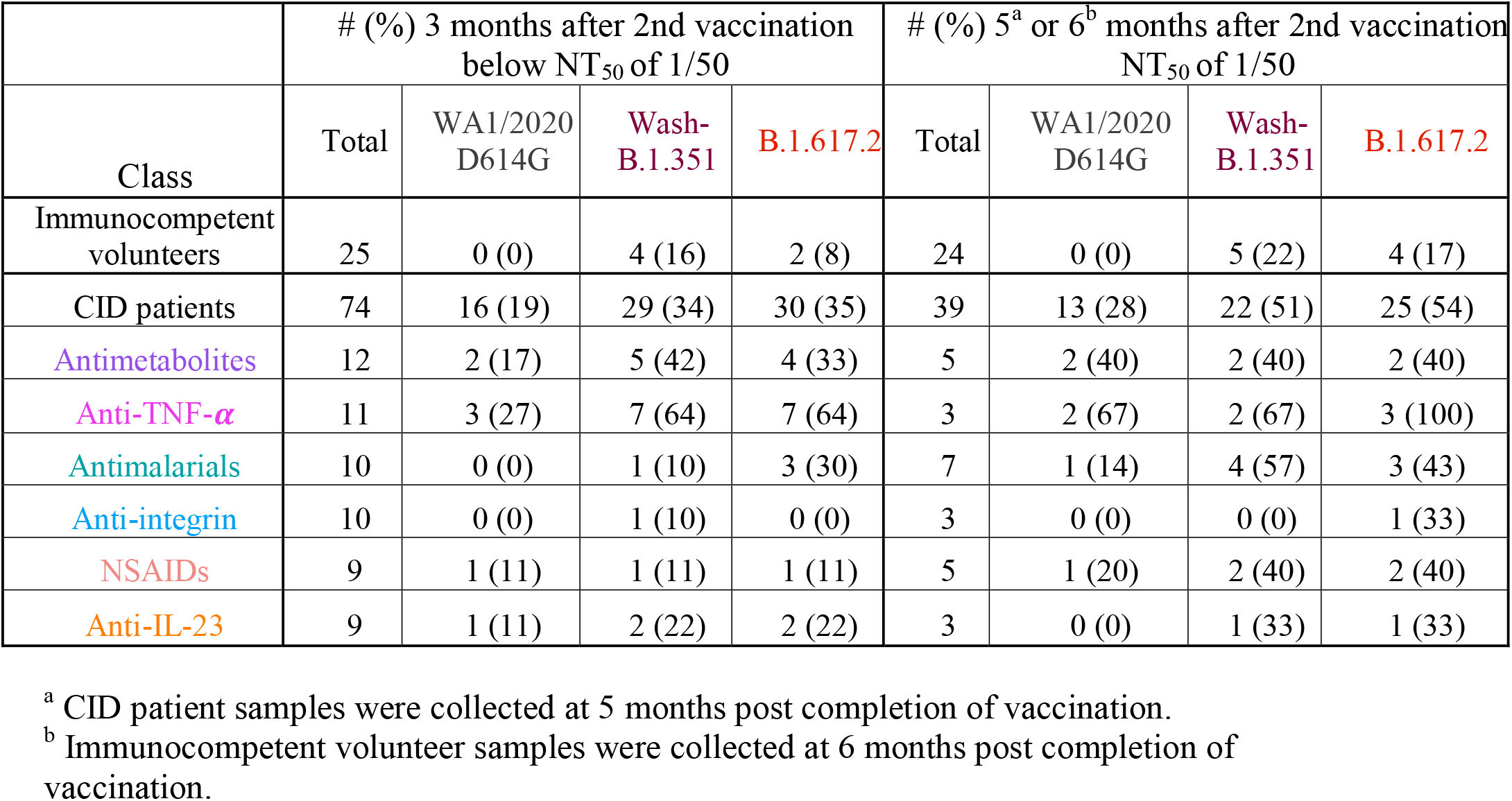
Number (and percentage) of individuals with neutralization titers (NT_50_) values below 1/50.

We separately grouped the individuals by disease and re-analyzed neutralization titers at three months after second vaccination. Some patient groups with certain diseases (*e*.*g*., ulcerative colitis, systemic lupus erythematosus, Sjogren’s syndrome, rheumatoid arthritis, and asthma) had neutralization titers that were similar to immunocompetent volunteers (**Fig 2A and S3**). In comparison, subjects with Crohn’s disease trended towards lower neutralization titers than healthy individuals (GMT: 239 (healthy) versus 159 (Crohn’s disease) [WA1/2020 D614G], *p* = 0.67; 117 versus 68 [Wash-B.1.351] *p* = 0.34; 109 versus 73 [B.1.617.2], *p* = 0.50) (**Fig 2A and S3**). Since TNFi is an important therapeutic target in Crohn’s disease, we stratified this group by treatment class. Crohn’s disease patients administered TNFi trended towards lower neutralizing titers than ones receiving other treatments (GMT: 107 (Crohn’s with TNFi) versus 198 (Crohn’s disease without TNFi) [WA1/2020 D614G], *p* = 0.27; 46 versus 85 [Wash-B.1.351] *p* = 0.30; 49 versus 93 [B.1.617.2], *p* = 0.26) (**Fig S3**). Compared to 34% of all CID patients at 3 months after second vaccination that had a neutralization titer of less than 1/50 against B.1.617.2, 50% of Crohn’s disease patients fell below this threshold (**Fig S3**). Moreover, 63% of Crohn’s disease patients receiving TNFi had neutralization titers against B.1.617.2 below the 1/50 cut-off compared to 43% of those on other treatment regimens (**Fig S3**). Thus, while Crohn’s disease patients had lower neutralizing titers after BNT162b2 mRNA vaccination than other CID patients, this effect appears confounded by TNFi use. Indeed, multivariable regression analysis indicated that the lower neutralization titers against B.1.617.2 were associated TNFi treatment and not Crohn’s disease (**Table 3**).

**Table 3.**
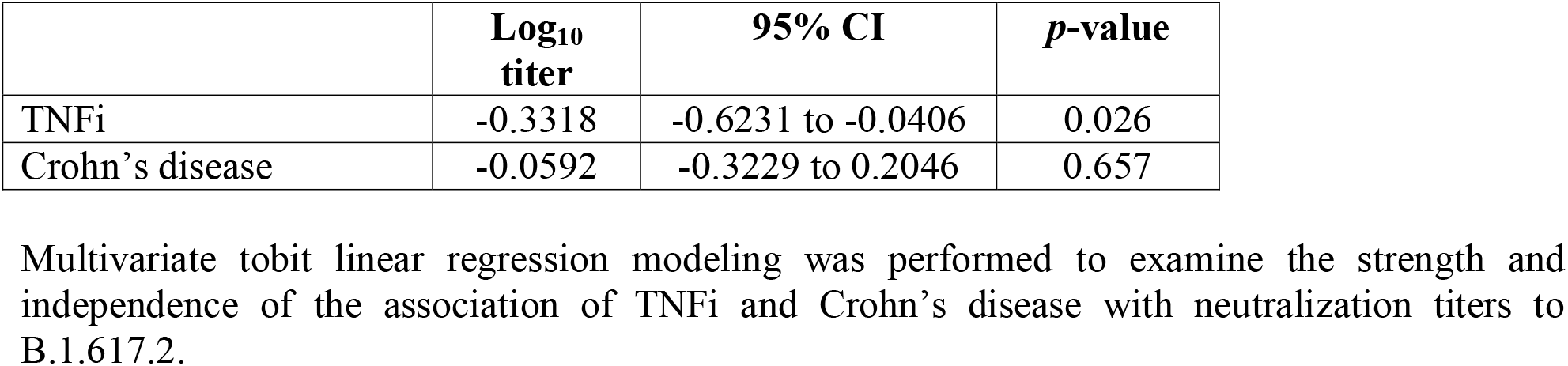
Regression analysis of effects on serum neutralization titers against B.1.617.2 infection.

Recent studies have shown that the Fc effector functions of antibodies can contribute to protection against SARS-CoV-2 infection and disease (Chan et al., 2021; Schäfer et al., 2021; Shiakolas et al., 2021; Winkler et al., 2021; Yamin et al., 2021; Zohar et al., 2020). To address whether our immunized CID patients had distinct Fc effector function profiles, we analyzed their SARS-CoV-2 specific antibodies in sera for IgG subclass distribution and C1q and Fcγ receptor (FcγR) binding. We focused our analyses on groups with at least n = 5: anti-integrin inhibitors, antimetabolites, TNFi, antimalarial agents, and immunocompetent volunteers, and measured responses against Wuhan-1 D614G, B.1.617.2, and B.1.351 spike proteins (**Fig 3** and **S4**). There were no differences in levels of IgG1, IgG3, and IgM against SARS-CoV-2 spike proteins, and C1q or FcγR (FcγR2A, FcγR2B, or FcγR3A) binding (**Fig 3** and **S4**). However, compared to immunocompetent volunteers, patients treated with TNFi had decreased anti-SARS-CoV-2 IgG, IgG2, and IgG4 levels against Wuhan-1 D614G (**Fig 3A**), B.1.617.2 (**Fig 3B**), and B.1.351 (**Fig S4**) whereas all other groups had no significant difference. We also assessed for antibody-mediated innate immune effector functions. While all groups showed similar antibody-dependent neutrophil phagocytosis (ADNP) (**Fig 3C-D** and **S4**), patients receiving TNFi had decreased antibody-dependent cellular phagocytosis (ADCP) against B.1.617.2 (**Fig 3D**) and B.1.351 (**Fig S3**). For unexplained reasons, patients receiving anti-integrin inhibitors had enhanced ADCP against Wuhan-1 D614G (**Fig 3C**) and antibody-dependent complement fixation (ADCD) against B.1.617.2 (**Fig 3D**) and B.1.351 (**Fig S4**). When we combined the data, only patients receiving TNFi showed substantive decreases in antibody effector functions at three months after second vaccination (**Fig 3E**).

**Figure 3.**
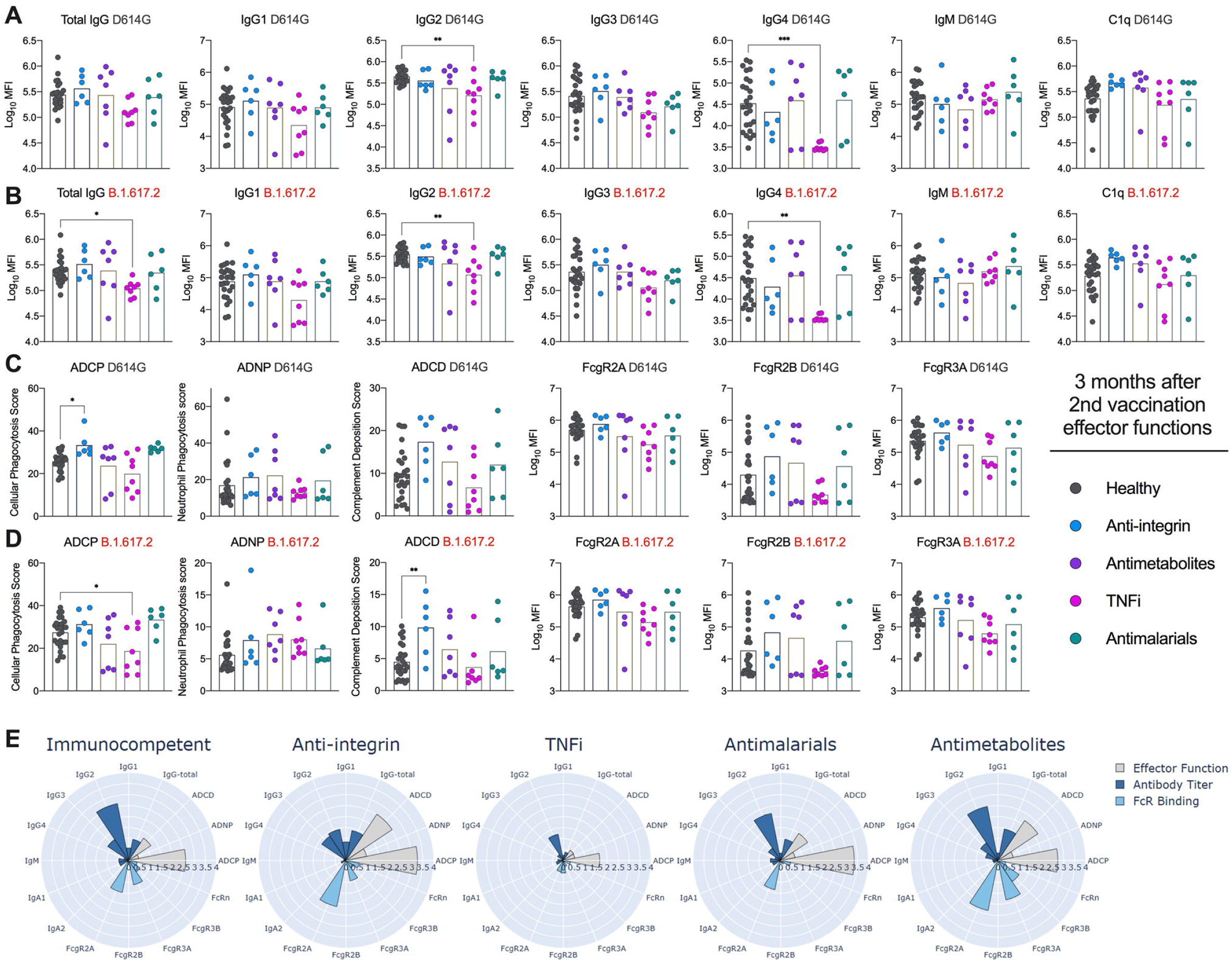
Effector functions against SARS-CoV-2 variant viruses at three months after second vaccination. Serum from humans at three months after second vaccination with BNT162b2 mRNA vaccine were assayed for (**A-B**) Total IgG, IgG subclasses (IgG1, IgG2, IgG3, IgG4, IgM), and C1q binding or (**C-D**) antibody-dependent cellular phagocytosis (ADCP), antibody-dependent neutrophil phagocytosis (ADNP), antibody-dependent complement deposition (ADCD), or FcγR (FcγR2A, FcγR2B, or FcγR3A) binding as measured by Luminex. Reponses were measured against (**A, C**) Wuhan-1 D614G or (**B, D**) B.1.617.2. Individuals were grouped as immunocompetent volunteers (n = 25) or subdivided by immunosuppressive drug class: TNFi (n = 8), antimetabolites (n = 7), antimalarials (n = 6), or anti-integrin inhibitors (n = 5). One-way ANOVA with Dunnett’s post-test; * *p* < 0.05; ** *p* < 0.01; *** *p* < 0.001. (**E**) Composite polar plots depicting shifted z-score only median of antibody titer, FcγR binding, and antibody function against Wuhan-1 D614G, B.1.351, and B.1.617.2 for each group.

As serum antibody titers generated by the Pfizer BNT162b2 vaccine wane over time(Israel et al., 2021; Thomas et al., 2021), we also assessed neutralizing activity at five months after immunization in CID patients (n = 39); for comparison, we used the later, six-month time point from our immunocompetent cohort (n =24) since a five-month time point was not sampled. Sera from CID patients trended toward lower neutralizing activity against all three viruses than immunocompetent volunteers (GMT: 178 (immunocompetent) versus 117 (CID patients) [WA1/2020 D614G], *P* = 0.77; 80 versus 61 [Wash-B.1.351], *P* = 0.53; 102 versus 59 [B.1.617.2], *P* = 0.18). Sera from both patient and immunocompetent subject groups also were less efficient at neutralizing Wash-B.1.351 and B.1.617.2 than WA1/2020 D614G (**Fig 4**). While there were limited numbers of patients in each drug group at this time point, the trend of decreased inhibitory activity against Wash-B.1.351 and B.1.617.2 was seen in patients receiving TNFi, anti-integrin inhibitors, NSAIDs, anti-IL-23 inhibitor, antimetabolites, antimalarial agents, BCDT, Nrf2 activator, SSZ, and anti-BLys) (**Fig 4 and S5**). Whereas only 4 of 24 (17%) immunocompetent volunteers had neutralization titers below 1/50 at the six-month time point against B.1.617.2, 25 of 39 (54%) of CID patients fell below this postulated protective threshold at the five-month time point (**Table 2**). Patients treated with antimetabolites (40%), TNFi (100%), antimalarial agents (43%), anti-integrin inhibitors (33%), NSAIDs (33%), and anti-IL-23 inhibitors (33%) had higher percentages of individuals than healthy volunteers below the 1/50 cut-off against B.1.617.2 (**Table 2**).

**Figure 4.**
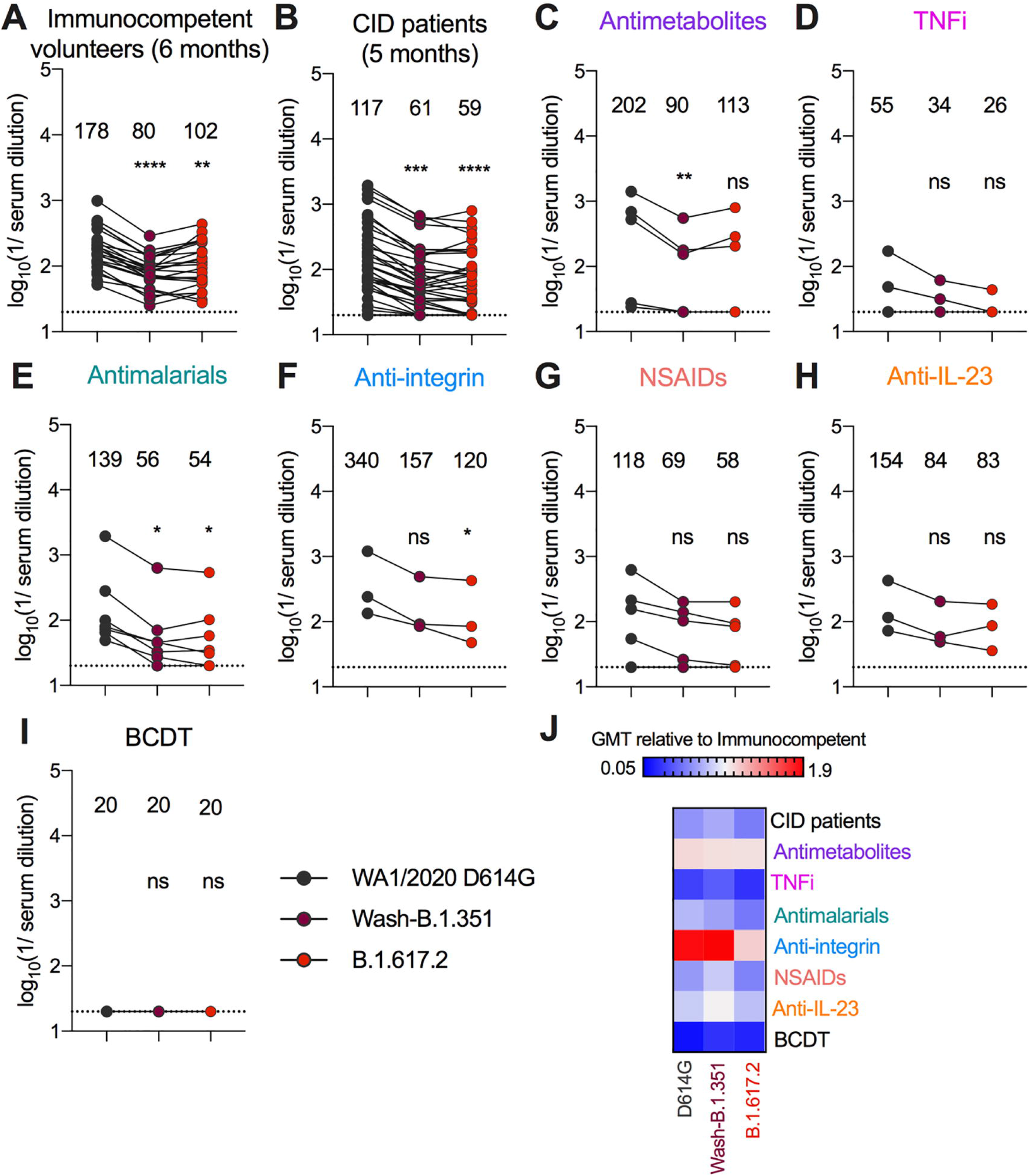
Serum neutralization titers of CID patients against SARS-CoV-2 variant viruses at five months after second vaccination. (**A-H**) Paired analyses of neutralization (NT_50_) titers in serum measured by FRNT against fully infectious SARS-CoV-2 strains from humans at five (CID patients) or six (immunocompetent volunteers) months after second vaccination with BNT162b2 mRNA vaccine; different time points were used based on variability of the separate study designs and availability of samples. Individuals were grouped as (**A**) immunocompetent volunteers (n = 24) (**B**) CID patients (n = 39) or subdivided by immunosuppressive drug class (**C**) antimetabolites (n = 5), (**D**) TNFi (n = 3), (**E**) antimalarials (n = 7), (**F**) anti-integrin inhibitors (n = 3), (**G**) NSAIDs (n = 5), (**H**) anti-IL-23 inhibitors (n = 3) or **(I)** B cell depletion therapy (BCDT) (n = 3). GMT values are shown on graph. Dotted line represents limit of detection of the assay. One way ANOVA with Dunn’s post-test; * *p* < 0.05; ** *p* < 0.01; *** *p* < 0.001; **** *p* < 0.0001. (**J**) Heat map of GMT values relative to healthy volunteer GMT values for each SARS-CoV-2 spike protein. Blue, reduction; red, increase.

## DISCUSSION

In this study, we evaluated the functional antibody responses after immunization with the Pfizer BNT162b2 mRNA vaccine against historical and emerging SARS-CoV-2 strains in a cohort of adult CID patients with a range of diagnoses and treatment interventions. These results were compared to a separate cohort of similarly vaccinated immunocompetent adults (Turner et al., 2021) and showed consistently lower serum antibody neutralizing titers in most CID patients, with a substantial fraction falling below an estimated 1/50 cutoff against B.1.617.2 (Delta) that has been proposed as a correlate of protection (Khoury et al., 2021). Subgroup analysis suggested that individuals on TNFi had lower inhibitory titers than other therapeutic groups. Thus, these patients might be at greatest risk for breakthrough infections, especially with VOC. Similarly, in studies that evaluated Fc effector function of serum antibodies, those receiving TNFi showed greater decreases in antibody effector functions, providing a second possible mechanism for risk of vaccine failure in these populations.

Our findings corroborate studies that report an association between patients with CID, including those on TNFi, and reduced antibody responses after vaccination (Alexander et al., 2021; Deepak et al., 2021; Fiorino et al., 2012). Poor seroconversion in these patients has been described after vaccination against hepatitis A (Park et al., 2014), hepatitis B (Haykir Solay and Eser, 2019; Pratt et al., 2018), and influenza (Cullen et al., 2012; Hua et al., 2014; Shirai et al., 2018) viruses which may be potentiated by TNFi and other immunosuppressants (Andrisani et al., 2013). Furthermore, a specific effect of TNFi on dampening antibody responses in IBD patients has been observed in those who recovered from SARS-CoV-2 infection (Dailey et al., 2021; Kennedy et al., 2021) or after vaccination (Edelman-Klapper et al., 2021). While we did observe that patients with Crohn’s disease had reduced neutralization titers against B.1.617.2, this was likely due to the use of TNFi rather than disease state itself. As TNF-α contributes to secondary lymphoid organ (B cell follicles or lymph nodes) development and signaling (Fu and Chaplin, 1999; Murphy and Weaver, 2016), TNFi may interfere with germinal center reactions and induction of optimal humoral immune responses.

The importance of Fc effector functions for antibody efficacy *in vivo* against SARS-CoV-2 has been illustrated with several NTD and RBD mAbs in animal models of infection (Schäfer et al., 2021; Suryadevara et al., 2021; Ullah et al., 2021; Winkler et al., 2021; Yamin et al., 2021). Consistent with these results, differences in antibody effector functions in serum also are associated with distinct levels of protection in the upper and lower respiratory tract of non-human primates immunized with a recombinant SARS-CoV-2 spike glycoprotein (NVX-CoV2373) (Gorman et al., 2021). Moreover, the BNT162b2 and mRNA1273 mRNA vaccines appear to induce different functional profiles in humans with higher RBD- and NTD-specific IgA, as well as functional antibodies (ADNP and ADNK) seen in mRNA-1273 vaccine recipients (Kaplonek et al., 2021). Thus, and independent of the lower neutralizing antibody levels observed in the sera of CID patients treated with TNFi, diminished Fc effector function profiles also could contribute to a higher frequency of breakthrough infections in this and other immunosuppressed vaccinated populations (Hall et al., 2021; Kamar et al., 2021; Qin et al., 2021).

The lower serological responses to the BNT162b2 vaccine seen in many of the CID patients in our cohort appears worse against the two viruses with spike genes of B.1.351 and B.1.617.2. This was not unexpected given that the evolution of more transmissible SARS-CoV-2 VOC with substitutions in the spike protein impacts antibody binding. Indeed, neutralization by vaccine-induced sera is diminished against variants expressing mutations in the spike gene at positions L452, E484, and elsewhere (Chen et al., 2021b; McCallum et al., 2021; Tada et al., 2021; Wang et al., 2021a; Wang et al., 2021b; Wibmer et al., 2021). Moreover, several vaccines have shown reduced ability to prevent symptomatic infection caused by the B.1.351 and B.1.617.2 variants in humans (Lopez Bernal et al., 2021; Madhi et al., 2021; Sadoff et al., 2021; Shinde et al., 2021). The lower functional (neutralizing and Fc effector functions) antibody responses seen in immunized immunosuppressed CID patients combined with inherently less inhibitory activity against emerging variants is consistent with the recent recommendation to administer an additional dose of vaccine to these at-risk populations. In addition, our data suggest that those receiving TNFi in particular should be further prioritized for an added vaccine dose and may require additional considerations such as temporary holding of immunosuppressive medication to optimize induction of protective immune responses.

### Limitations of study

We note several limitations in our study. (1) Due to the diversity of immunosuppressants used, the number in each treatment subgroup is relatively small, limiting the power of the statistical analysis; (2) The cohort we used was a subset of a larger CID cohort (Deepak et al., 2021) that included vaccinated patients receiving multiple immunosuppressive treatment modalities. Only those on monotherapy regimens were evaluated in our study to eliminate confounding by use of concomitant immunosuppressive agents; (3) We grouped subjects by treatment intervention but due to the small numbers did not account for underlying disease severity nor diagnosis, which independently could impact immune responses; (4) Our study focused on subjects immunized with the Pfizer BNT162b2 mRNA vaccine. Separate studies are needed with other vaccine platforms; (5) Our studies focused on the impact of immunosuppression on serum antibody responses after vaccination and did not account for T cell and anamnestic responses, which also may confer protection; and (6) The small sample size precluded assessment of comorbidities, age, and sex on humoral responses.

From a cohort of 74 patients with chronic inflammatory diseases on different immunosuppressive therapy, we observed a clear trend towards lower antibody neutralizing and Fc effector function responses after two doses of Pfizer BNT162b2 mRNA vaccination in those receiving TNFi. As these responses are even lower against emerging VOC, boosting and functional monitoring of immunity will be important in these individuals. Future studies are warranted to understand and improve SARS-CoV-2 immunity in this and other immunologically vulnerable populations.

## Data Availability

All data supporting the findings of this study are available within the paper and are available from the corresponding author upon request.

## ACKNOWLEDGEMENTS

This study was supported by grants and contracts from NIH (R01 AI157155, R01AI151178, HHSN75N93019C00074, NIAID Centers of Excellence for Influenza Research and Response (CEIRR) contracts HHSN272201400008C and 75N93021C00014, and the Collaborative Influenza Vaccine Innovation Centers (CIVIC) contract 75N93019C00051). The Alter laboratory was supported by the Ragon Institute, the Massachusetts Consortium on Pathogen Readiness (MassCPR), the NIH (3R37AI080289-11S1, R01AI146785, U19AI42790, U19AI135995, U19AI42790, 1U01CA260476, CIVIC75N93019C00052), the Gates foundation Global Health Vaccine Accelerator Platform funding (OPP1146996 and INV-001650), and the Musk Foundation. M.P. was supported by the Scientist Development Award from the Rheumatology Research Foundation. P.D. is supported by a Junior Faculty Development Award from the American College of Gastroenterology and IBD Plexus of the Crohn’s & Colitis Foundation. A.H.J.K. is supported by the Rheumatology Research Foundation, NIH/NIAMS P30 AR073752, and PCORI SDM2017C28224. G.F.W. was supported by grant funding from the NIH (R01 NS106289) and the NMSS (RG-1802-30253). The COVaRiPAD study was supported by The Leona M. and Harry B. Helmsley Charitable Trust, Washington University Digestive Disease Research Core Center (NIDDK P30DK052574), Washington University Rheumatic Diseases Research Resource-Based Center (NIAMS P30AR073752), the Judy Miniace Research Fund for the Washington University Lupus Clinic, the Siteman Cancer Center grant P30CA091842 from the NIH/NCI, and the Washington University Institute of Clinical and Translational Sciences grant UL1TR002345 from the NIH/NCATS. We thank Richard Webby and Pei-Yong Shi for some of the viruses used in this study and Kimberly E. Taylor at University of California, San Francisco for statistical support.

## AUTHOR CONTRIBUTIONS

R.E.C. performed and analyzed neutralization assays. M.J.G., D.Y., and D.Y.Z. performed and analyzed effector function analyses. J.M.C. performed and analyzed ELISA data. L.D. and S.A.H. performed and analyzed next generation sequencing of viral stocks. P.D., M.P., R.M.P., J.A.O’H., S.C., G.F.W., A.H.E., and A.H.J.K. designed the clinical studies and provided human samples. S.B., W.K., and J.S.T. processed clinical samples. P.D., D.A.L., F.K., G.A., A.H.E., A.H.J.K., and M.S.D. obtained funding. G.A. and M.S.D. supervised the research. R.E.C. and M.S.D. wrote the initial draft, with the other authors providing editorial comments.

## SUPPLEMENTAL FIGURE LEGENDS

**Figure S1. Serum IgG titers against SARS-CoV-2 variant spike proteins at three months after second vaccination, small n**. Paired analyses of spike-specific endpoint IgG serum titers measured by ELISA from humans at three months after second vaccination with BNT162b2 mRNA vaccine. Individuals were grouped by immunosuppressive drug class: anti-IL-23 inhibitors, B cell depletion therapy (BCDT), Bruton’s tyrosine kinase inhibitor (BTKi), nuclear factor erythroid-2-related factor 2 (Nrf2) activator, sulfasalazine (SSZ), systemic steroid, anti-B lymphocyte stimulator (anti-BLyS), and sphingosine 1-phosphate receptor modulator (S1PR mod). GMT values are shown on graph. Dotted line represents limit of detection of the assay.

**Figure S2. Serum neutralization titers against SARS-CoV-2 variant viruses at three months after second vaccination, small n**. Paired analyses of neutralization (NT_50_) titers in serum measured by FRNT from humans at three months after second vaccination with BNT162b2 mRNA vaccine. Individuals were grouped by immunosuppressive drug class: B cell depletion therapy (BCDT), Bruton’s tyrosine kinase inhibitor (BTKi), nuclear factor erythroid-2-related factor 2 (Nrf2) activator, sulfasalazine (SSZ), systemic steroid, anti-B lymphocyte stimulator (anti-BLyS), and sphingosine 1-phosphate receptor modulator (S1PR mod). GMT values are shown on graph. Dotted line represents limit of detection of the assay.

**Figure S3. Serum neutralization titers against SARS-CoV-2 variant viruses at three months after second vaccination by CID classification**. (***Left***) Paired analyses of neutralization (NT_50_) titers in serum measured by FRNT from humans at three months after second vaccination with BNT162b2 mRNA vaccine. Individuals were grouped by CID diagnosis: Crohn’s disease, ulcerative colitis, systemic lupus erythematosus, Sjogren’s syndrome, rheumatoid arthritis, asthma, and multiple sclerosis. Crohn’s disease patients were further divided by treatment with or without anti-TNF-α inhibitors. GMT values are shown on graph. Dotted line represents limit of detection of the assay. One way ANOVA with Dunn’s post-test; * *p* < 0.05; ** *p* < 0.01; *** *p* < 0.001; **** *p* < 0.0001. (***Right***) Table with number (or percentage) of patients with NT_50_ values below 1/50 for each group against WA1/2020 D614G, Wash-B.1.351, and B.1.617.2.

**Figure S4. Effector functions against SARS-CoV-2 variant viruses at three months after second vaccination against B.1.351**. Serum from humans at three months after second vaccination with BNT162b2 mRNA vaccine were assayed for Total IgG, IgG subclasses (IgG1, IgG2, IgG3, IgG4, IgM), C1q binding, antibody-dependent cellular phagocytosis (ADCP), antibody-dependent neutrophil phagocytosis (ADNP), antibody-dependent complement deposition (ADCD), or FcγR (FcγR2A, FcγR2B, or FcγR3A) binding as measured by Luminex. Reponses were measured against B.1.351. Individuals were grouped as immunocompetent volunteers (n = 25) or subdivided by immunosuppressive drug class: TNFi (n = 8), antimetabolites (n = 7), antimalarials (n = 6), or anti-integrin inhibitors (n = 5). One-way ANOVA with Dunnett’s post-test; * *p* < 0.05; ** *p* < 0.01.

**Figure S5. Serum neutralization titers of CID patients against SARS-CoV-2 variant viruses at five months after second vaccination, small n**. Paired analyses of neutralization (NT_50_) titers in serum measured by FRNT from humans at five months after second vaccination with BNT162b2 mRNA vaccine. Individuals were grouped by immunosuppressive drug class: nuclear factor erythroid-2-related factor 2 (Nrf2) activator, sulfasalazine (SSZ), anti-B lymphocyte stimulator (anti-BLyS), Bruton’s tyrosine kinase inhibitor (BTKi), systemic steroid, and sphingosine 1-phosphate receptor modulator (S1PR mod). GMT values are shown on graph. Dotted line represents limit of detection of the assay.

## STAR METHODS

### RESOURCE AVAILABLITY

#### Lead Contact

Further information and requests for resources and reagents should be directed to the Lead Contact, Michael S. Diamond.

#### Materials Availability

All requests for resources and reagents should be directed to the Lead Contact author. This includes mice and viruses. All reagents will be made available on request after completion of a Materials Transfer Agreement.

### EXPERIMENTAL MODEL AND SUBJECT DETAILS

#### Patient samples

CID patient samples were obtained from the COVaRiPAD longitudinal observational study as previously described (Deepak et al., 2021). Immunocompetent volunteer samples were obtained as previously described (Turner et al., 2021). All individuals were enrolled at Washington University School of Medicine in studies that had received Institutional Review Board approval (202012081 (WU368) and 202012084 (COVaRiPAD).

#### Cells

Vero-TMPRSS2 cells (Zang et al., 2020) were cultured at 37°C in Dulbecco’s Modified Eagle medium (DMEM) supplemented with 10% fetal bovine serum (FBS), 10□mM HEPES pH 7.3, 1□mM sodium pyruvate, 1× non-essential amino acids, and 100□U/ml of penicillin–streptomycin, and 5 μg/mL of blasticidin.

#### Viruses

The WA1/2020 D614G recombinant strain was obtained from an infectious cDNA clone of the 2019n-CoV/USA_WA1/2020 strain as described previously (Plante et al., 2021). The Beta (B.1.351) variant spike gene was introduced into the WA1/2020 backbone as described previously (Chen et al., 2021b). The B.1.617.2 isolate was obtained a gift from R. Webby (Memphis, TN). All viruses were passaged once in Vero-TMPRSS2 cells and subjected to next-generation sequencing after RNA extraction to confirm the introduction and stability of substitutions (**Table S1**). All virus experiments were performed in an approved Biosafety level 3 (BSL-3) facility.

### METHOD DETAILS

#### Enzyme-linked immunosorbent assay (ELISA)

Serum antibodies against SARS-CoV-2 spike (S) were measured as previously described (Amanat et al., 2020; Stadlbauer et al., 2020). Briefly, polystyrene 96-well plates (Immulon 4HBX; Thermo Fisher Scientific) were coated with 50□μL/well of PBS (pH□7.4) (Gibco) containing recombinant spike proteins (2□μg/mL) and incubated at 4°C overnight. On the next day, plates were washed with PBS-0.1% Tween 20 (PBS-T) using an automated plate washer (AquaMax 2000; Molecular Devices). Plates were blocked with 220□μL/well of PBS-T, 3% nonfat dry milk (AmericanBio) for 1 h. For serum and secondary antibody dilutions, a solution of PBS-T, 1% nonfat dry milk (AmericanBio) was used. Sera were serially diluted (3-fold) starting at a 1:100 dilution. Dilutions were added to the plates (100□μL/well) for 2 h at room temperature (RT). Plates were washed, and the secondary antibody IgG (whole molecule)-peroxidase antibody was added for 1 h at room temperature. Plates were washed, and the substrate *o*-phenylenediamine dihydrochloride (SigmaFast OPD; Sigma-Aldrich) was added (100□μL/well) and incubated for 10□min. The reaction was stopped by addition of 50□μL/well of a 3 M HCl solution (Thermo Fisher Scientific). Optical density (OD) was measured (490□nm) using a microplate reader (Synergy H1; BioTek). Analysis was performed using Prism 7 software (GraphPad), and values were reported as area under the curve (AUC).

#### Focus reduction neutralization test

Serial dilutions of mAbs or sera were incubated with 10^2^ focus-forming units (FFU) of different strains or variants of SARS-CoV-2 for 1 h at 37°C. Antibody-virus complexes were added to Vero-TMPRSS2 cell monolayers in 96-well plates and incubated at 37°C for 1 h. Subsequently, cells were overlaid with 1% (w/v) methylcellulose in MEM supplemented with 2% FBS. Plates were harvested 24 h later by removing overlays and fixed with 4% PFA in PBS for 20 min at room temperature. Plates were washed and sequentially incubated with an oligoclonal pool of SARS2-2, SARS2-11, SARS2-16, SARS2-31, SARS2-38, SARS2-57, and SARS2-71 (Liu et al., 2021d; VanBlargan et al.) anti-spike antibodies and HRP-conjugated goat anti-mouse IgG (Sigma, 12-349) in PBS supplemented with 0.1% saponin and 0.1% bovine serum albumin. SARS-CoV-2-infected cell foci were visualized using TrueBlue peroxidase substrate (KPL) and quantitated on an ImmunoSpot microanalyzer (Cellular Technologies).

#### Effector function antigens

Antigens used for Luminex based assays: SARS-CoV-2 D614G, Beta B.1.351, and Delta B.1.617.2 spike antigens all were kindly provided by Erica Ollmann Saphire (La Jolla Institute for Immunology).

#### Luminex profiling

Serum samples were analyzed by customized Luminex assay to quantify the relative concentration of antigen-specific antibody isotypes, subclasses, and Fcγ-receptor (FcγR) binding profiles, as previously described (Brown et al., 2017; Brown et al., 2012). Briefly, SARS-CoV-2 antigens were used to profile specific humoral immune responses. Antigens were coupled to magnetic Luminex beads (Luminex Corp) by carbodiimide-NHS ester-coupling (Thermo Fisher). Antigen-coupled microspheres were washed and incubated with plasma or serum samples at an appropriate sample dilution (1:5000 for IgG1 and all low affinity FcγR, and 1:200 for all other readouts) for 2 h at 37°C in 384-well plates (Greiner Bio-One). Unbound antibodies were washed away, and antigen-bound antibodies were detected by using a PE-coupled detection antibody for each subclass and isotype (IgG1, IgG3, IgA1, and IgM; Southern Biotech), and FcγR were fluorescently labeled with PE before addition to immune complexes (FcγR2a, FcγR3a; Duke Protein Production facility). After one hour of incubation, plates were washed, and flow cytometry was performed with an iQue (Intellicyt) and analyzed using IntelliCyt ForeCyt (v8.1). PE median fluorescent intensity (MFI) is reported as a readout for antigen-specific antibody titers.

#### Antibody-dependent complement deposition (ADCD)

Antibody-dependent complement deposition (ADCD) was conducted as previously described(Fischinger et al., 2019). Briefly, SARS-CoV-2 antigens were coupled to magnetic Luminex beads (Luminex Corp) by carbodiimide-NHS ester-coupling (Thermo Fisher). Coupled beads were incubated for 2 h at 37°C with serum samples (1:10 dilution) to form immune complexes and then washed to remove unbound immunoglobulins. To measure antibody-dependent deposition of C3, lyophilized guinea pig complement (Cedarlane) was diluted in gelatin veronal buffer with calcium and magnesium (GBV++) (Boston BioProducts) and added to immune complexes. Subsequently, C3 was detected with an anti-C3 fluorescein-conjugated goat IgG fraction detection antibody (Mpbio). Flow cytometry was performed 5 Laser LSR Fortessa Flow Cytometer and analyzed using FlowJo V10.7.1. ADCD was reported as the median of C3 deposition.

#### Antibody-dependent cellular (ADCP) and neutrophil (ADNP) phagocytosis

Antibody-dependent cellular phagocytosis (ADCP) and antibody-dependent neutrophil phagocytosis (ADNP) were conducted according to the previously described protocols (Butler et al., 2019; Karsten et al., 2019). In detail, SARS-CoV-2 antigens were biotinylated using EDC (Thermo Fisher) and Sulfo-NHS-LCLC biotin (Thermo Fisher) and coupled to yellow-green (505/515) fluorescent Neutravidinconjugated beads (Thermo Fisher), respectively. To form immune complexes, antigen-coupled beads were incubated for 2 h at 37°C with 1:100 diluted serum samples and then washed to remove unbound immunoglobulins. For ADCP, the immune complexes were incubated for 16–18 hours with THP-1 cells (1.25×10^5^ THP-1 cells/mL) and for ADNP for 1 hour with RBC-lyzed whole blood. Following the incubation, cells were fixed with 4% PFA. For ADNP, RBC-lyzed whole blood was washed, stained for CD66b^+^ (Biolegend) to identify neutrophils, and then fixed in 4% PFA. Flow cytometry was performed to identify the percentage of cells that had phagocytosed beads as well as the number of beads that had been phagocytosis (phagocytosis score = % positive cells × Median Fluorescent Intensity of positive cells/10000). Flow cytometry was performed with 5 Laser LSR Fortessa Flow Cytometer and analyzed using FlowJo V10.7.1.

### QUANTIFICATION AND STATISTICAL ANALYSIS

Statistical significance was assigned when *P* values were < 0.05 using Prism Version 8 (GraphPad). Specific tests (one-way ANOVA with Dunn’s or Dunnett’s post-test for multiple comparisons), number of subjects (n), geometric mean values, and comparison groups are indicated in the Figure legends. All data were graphed and analyzed in GraphPad Prism v8.4.3. The data used to generate **Fig 3e** was graphed and analyzed using Python version 3.8.5 and the ‘plotly’ package (Sievert, 2020). For each feature, data was first standardized by computing the Z-score, scaling values to zero mean and unit variance. The median resulting values are represented on each polar plot. All other data were graphed and analyzed in GraphPad Prism v8.4.3. Tobit linear regression was performed using Stata/MP 13.1, and the effects were refined to account for left-censoring of data below the limit of detection (LoD).

