## Supplementary figures and images for "Reduced antibody activity against SARS-CoV-2 B.1.617.2 Delta virus in serum of mRNA-vaccinated patients receiving TNF-α inhibitors"

### Figure S1

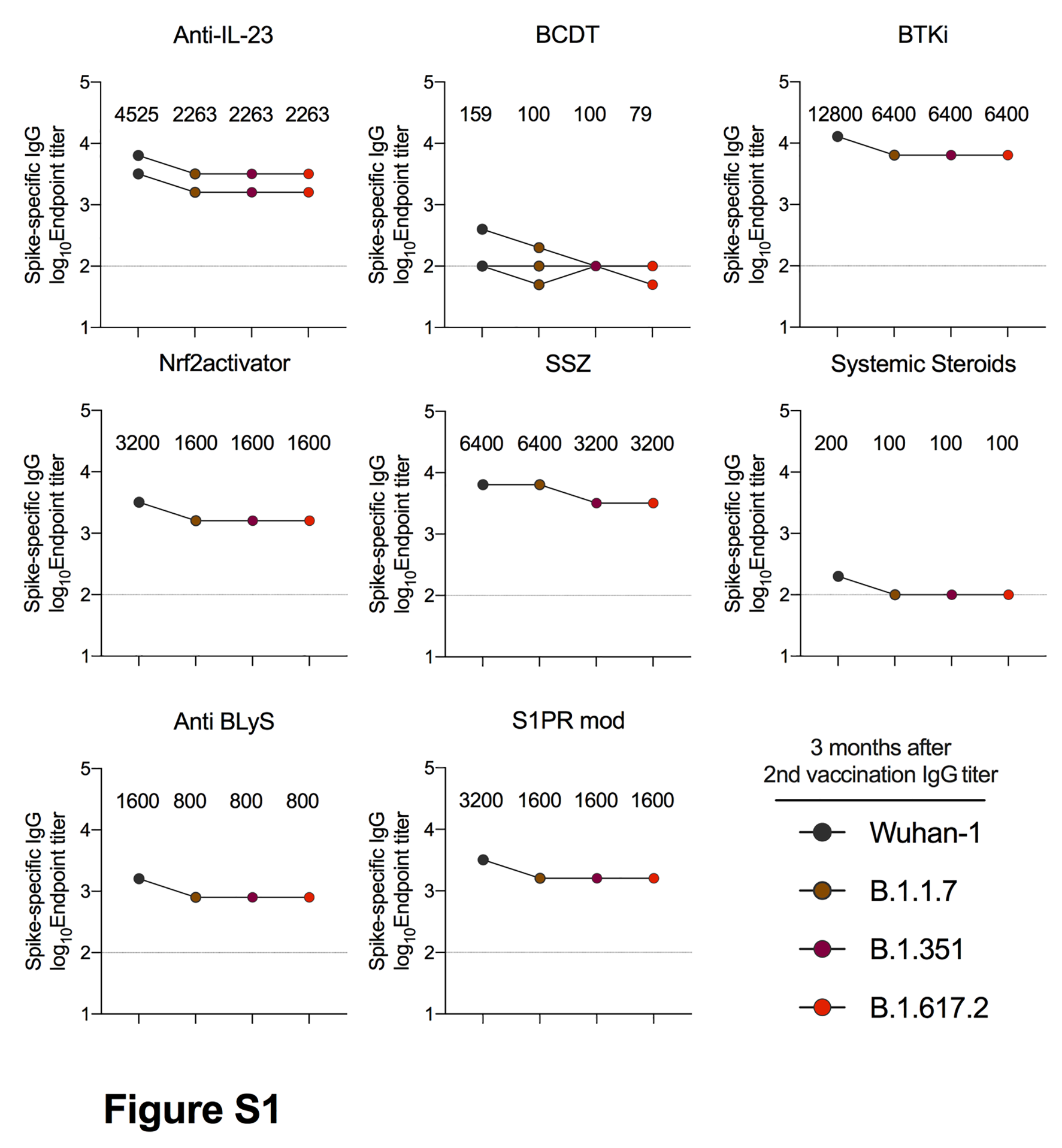

### Figure S2

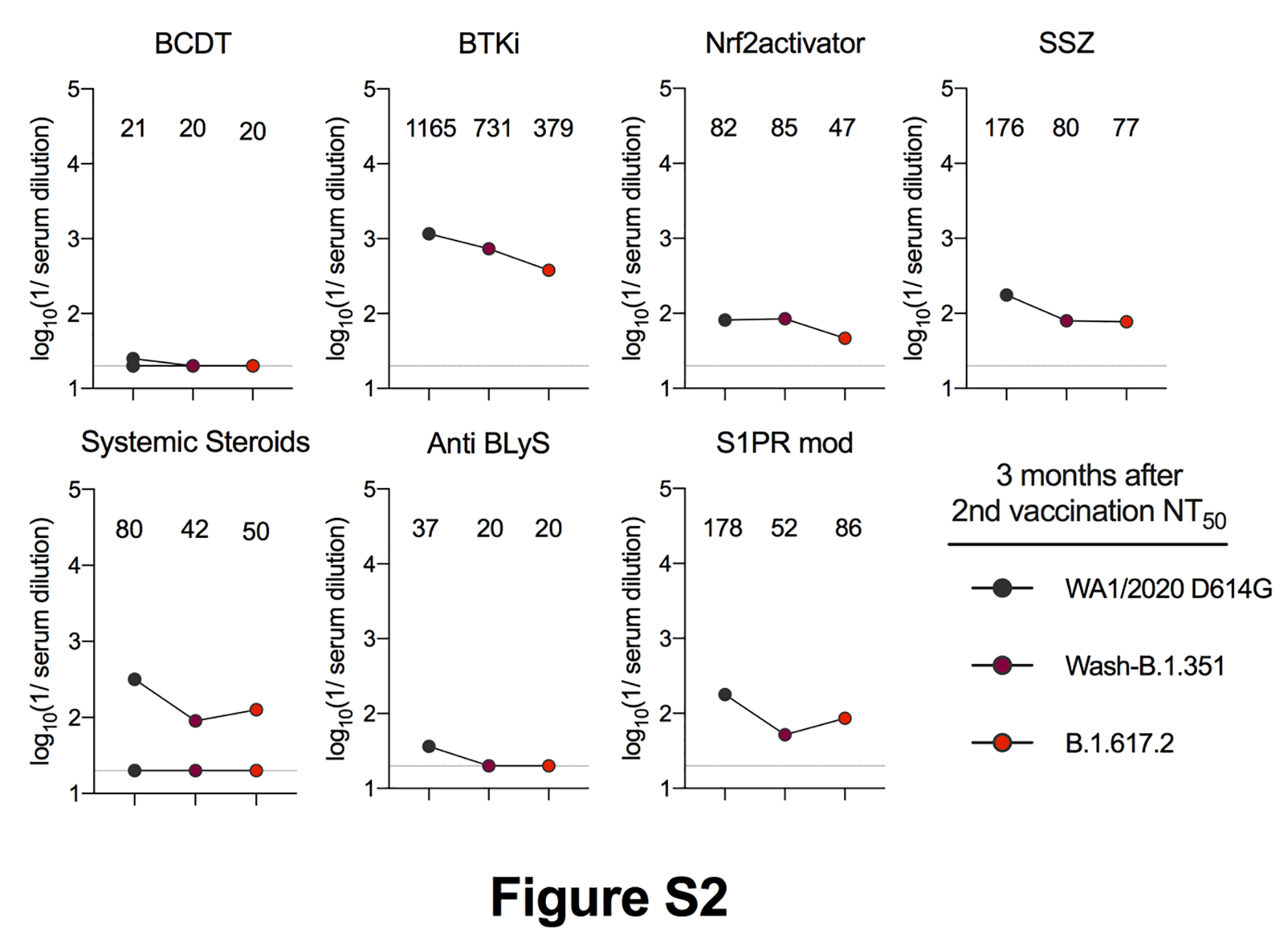

### Figure S3

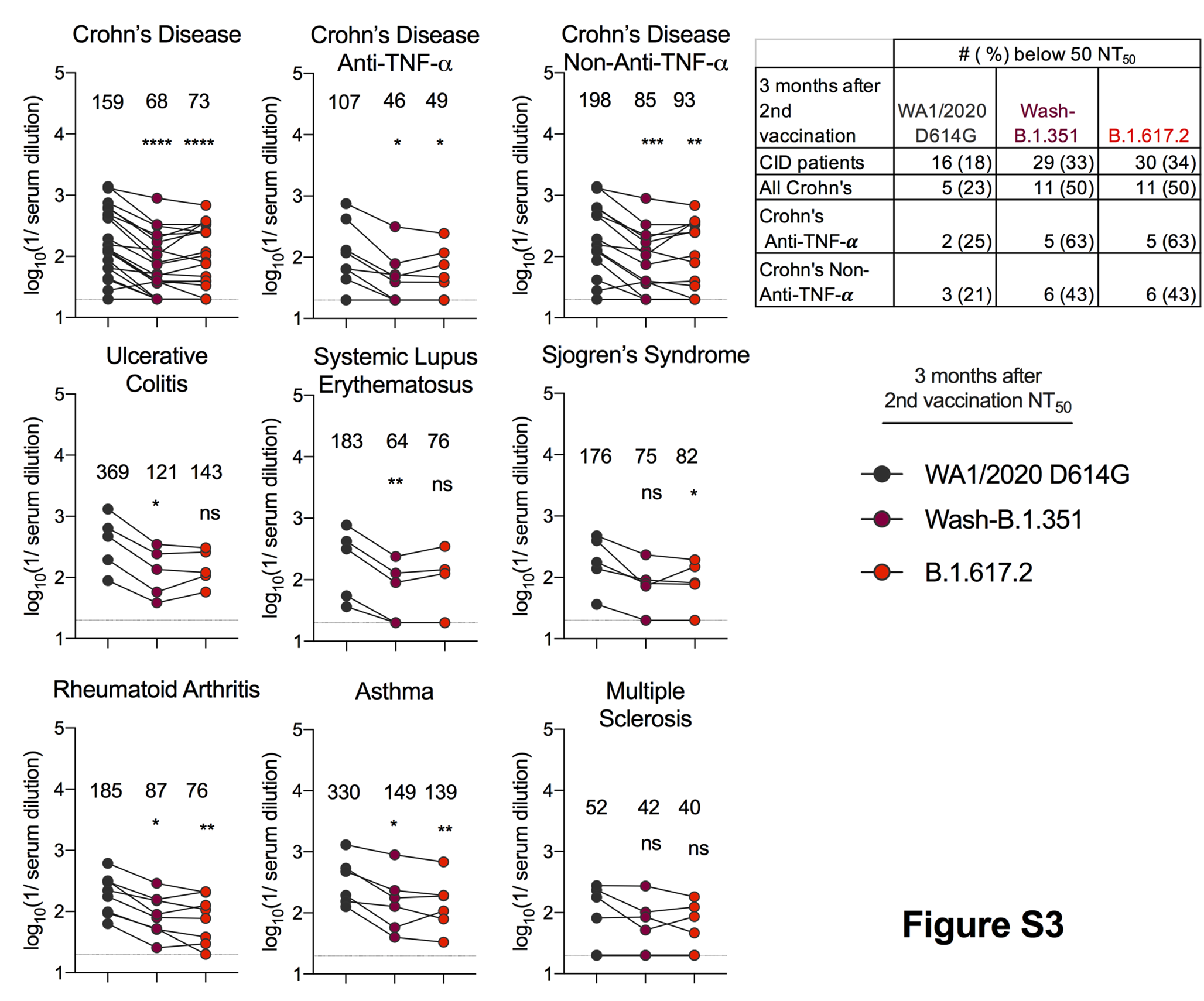

### Figure S4

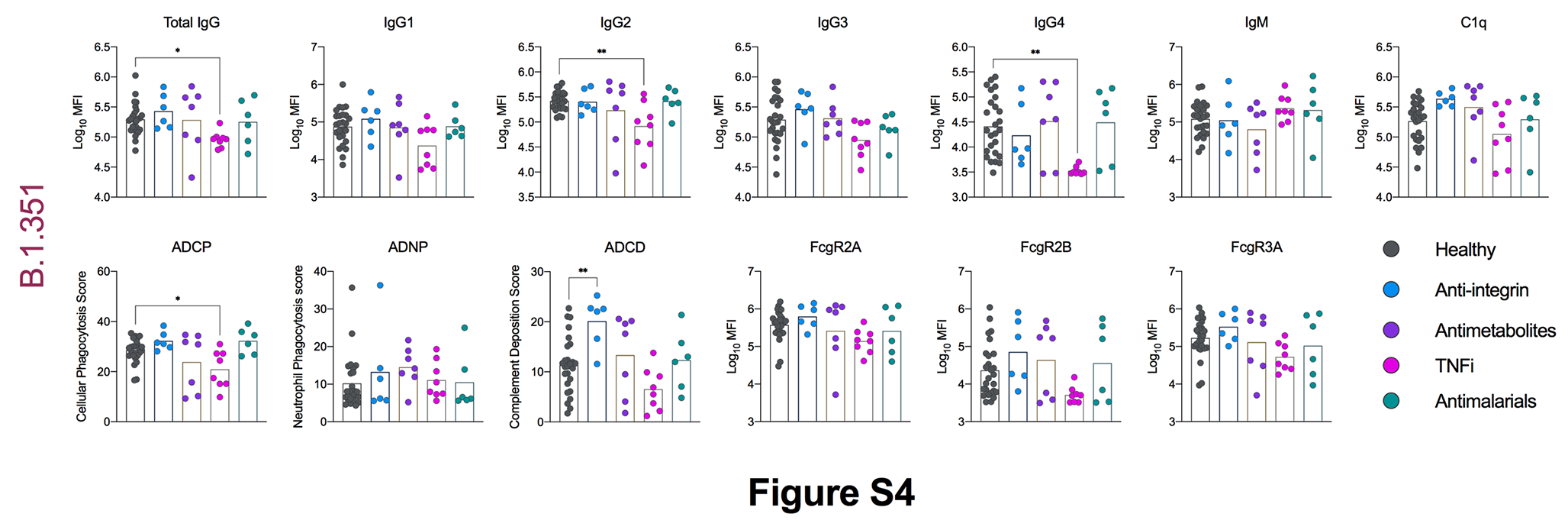

### Figure S5

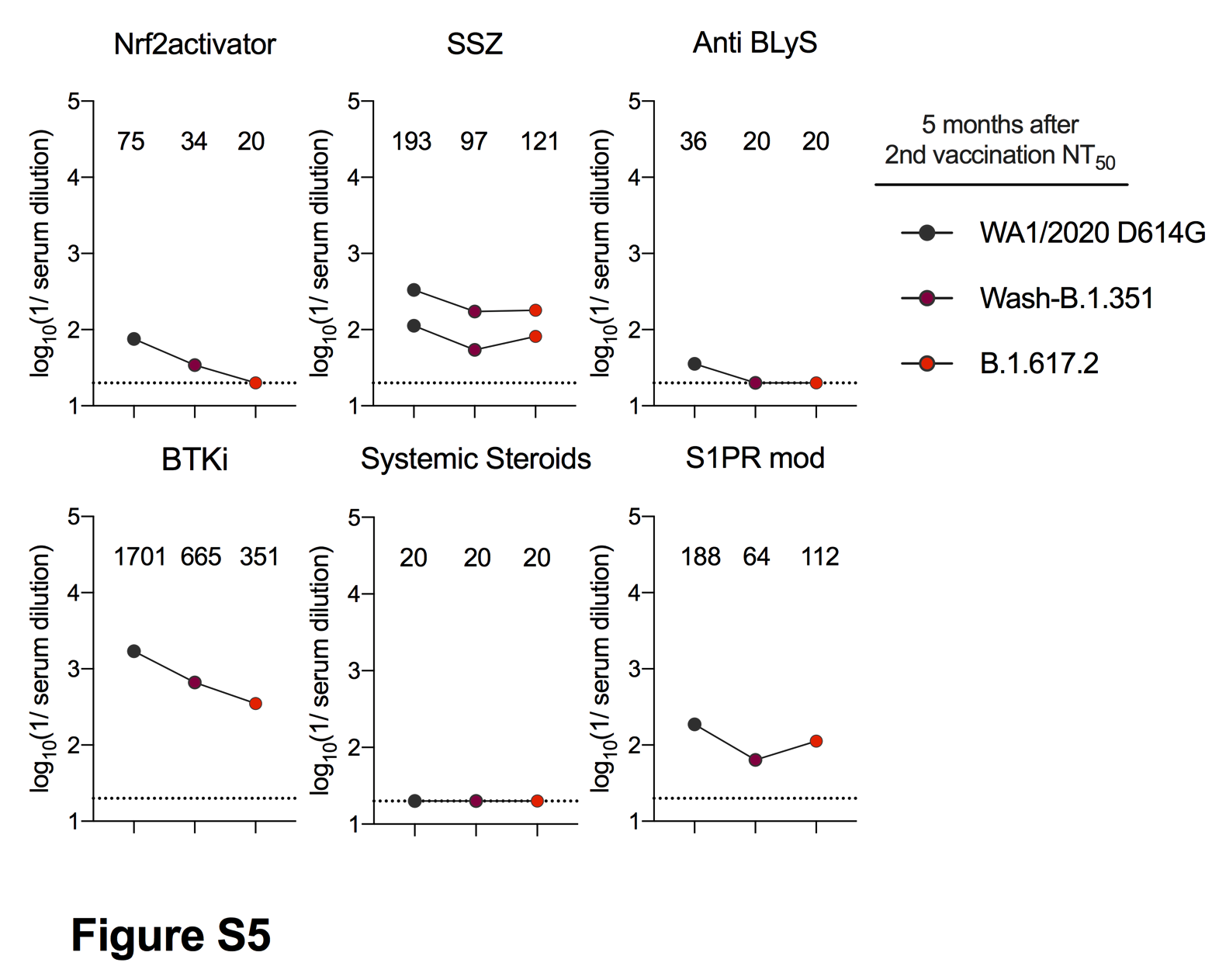
